# Baseline Serum Metabolites as Predictors of Teplizumab Response in Individuals with Type 1 Diabetes

**DOI:** 10.64898/2025.12.22.25342822

**Authors:** Elizabeth R. Flammer, Lauren E. Higdon, Srinath Sanda, Timothy J. Garrett, Heba M. Ismail

## Abstract

**Aims/hypothesis:** Immunotherapies such as Teplizumab can preserve residual beta cell function in individuals with newly diagnosed type 1 diabetes (T1D), but treatment response is variable. Currently, no biomarker exists to identify individuals most likely to benefit from immunotherapy. We believe that baseline serum metabolomic profiles can distinguish individuals who respond to treatment from nonresponders and predict therapeutic response.

**Methods:** Baseline serum samples from 41 individuals newly diagnosed with T1D enrolled in the AbATE trial (NCT00129259) were analyzed to identify metabolic predictors of response to Teplizumab therapy in the AbATE trial. Responders to Teplizumab, as per study protocol, were defined as individuals who exhibited less than a 40% decline in baseline C-peptide levels at 2 years after start of treatment. We analyzed baseline serum samples using a semi-targeted metabolomics approach via liquid chromatography–high-resolution tandem mass spectrometry. Metabolites that were significantly different between responders and nonresponders were identified (P < 0.05), and the significant metabolites were used to train a supervised Random Forest model to predict treatment response. Model performance was evaluated using a 70/30 training/testing split, 5-fold cross-validation, bootstrap resampling (1,000 iterations), and permutation testing (1,000 permutations).

**Results:** We identified 15 significantly different metabolites at baseline between responders and nonresponders (P < 0.05). These metabolites included amino acids and their derivatives, tricarboxylic acid (TCA) cycle intermediates, and microbially derived metabolites. At baseline, responders exhibited higher levels of TCA cycle metabolites, amino acid derivatives, and microbial metabolites, whereas nonresponders showed elevated levels of glutamate and acylcarnitines. The Random Forest classifier achieved an accuracy of 0.769 and an area under the receiver operating characteristic curve (AUC) of 0.881 in the test dataset. Cross-validation yielded a mean AUC of 0.856 (SD 0.156; 95% CI 0.719–0.992). Bootstrap analysis produced a test AUC 95% CI of 0.619–1.000, and permutation testing confirmed significance (p = 0.012).

**Conclusions/interpretation:** Baseline serum metabolomic signatures can predict responders to Teplizumab with high accuracy. This could potentially be applicable when considering other immunotherapies in preventative efforts in T1D.

**Trial registration:** ClinicalTrials.gov NCT00129259.

**Research in Context:** *What is already known about this subject?:*

- Teplizumab can delay beta cell decline in individuals with newly diagnosed T1D, but treatment response varies.
- No validated biomarkers currently exist to predict which individuals will respond to immunotherapy.
- Metabolomic profiling has shown potential for identifying metabolic signatures associated with disease progression and immune activity in T1D.

*What is the key question?:*

- Can baseline serum metabolomic profiles predict which individuals with newly diagnosed T1D will respond to Teplizumab therapy?

*What are the new findings?:*

- Fifteen baseline metabolites differed significantly between responders and nonresponders, including amino acid derivatives, tricarboxylic acid cycle intermediates, and microbially derived metabolites.
- Responders exhibited metabolic signatures consistent with preserved beta cell function and enhanced mitochondrial and immune-regulatory activity.
- A Random Forest model developed using these metabolites accurately predicted treatment response (AUC 0.881), demonstrating strong predictive potential.

*How might this impact on clinical practice in the foreseeable future?:*

- Baseline metabolomic profiling could support personalized treatment strategies by identifying individuals most likely to benefit from treatment with Teplizumab or other immunotherapies.

## Introduction

Type 1 diabetes (T1D) is an autoimmune disease that affects both children and adults, characterized by T-cell mediated destruction of insulin-producing pancreatic beta cells, ultimately resulting in a lifelong dependence on exogenous insulin therapy. The disease develops gradually over months to years and is often preceded by the appearance of circulating islet autoantibodies [1]. Prior to clinical diagnosis, significant β-cell loss has typically occurred, as indicated by reduced levels of C-peptide, a marker of endogenous insulin secretion [2]. C-peptide is secreted in equimolar amounts with insulin from pancreatic beta cells and serves as a reliable biomarker of residual beta cell function. Following diagnosis, C-peptide levels generally decline rapidly, particularly within the first five years, reflecting progressive autoimmune destruction of beta cells [2].

It has been shown that immune-based therapies can preserve residual beta cell function in people with T1D. However, therapeutic response varies among individuals, with some showing minimal or no improvement following treatment [3]. Teplizumab, an Fc receptor–nonbinding anti-CD3 monoclonal antibody, has been developed to mitigate beta cell loss shown by reducing the decline in C-peptide levels in patients with T1D [3]. The Autoimmunity-Blocking Antibody for Tolerance in Recently Diagnosed Type 1 Diabetes (AbATE) trial (ClinicalTrials.gov identifier: NCT00129259) evaluated individual responses to Teplizumab treatment to determine whether treatment could slow C-peptide decline over two years following disease onset [3, 4]. Newly diagnosed individuals were treated with Teplizumab for 2 weeks (median cumulative dose 11.6 mg, diluted in normal saline and administered intravenously at diagnosis) and participants who demonstrated detectable C-peptide responses during mixed-meal tolerance tests received a second course one year later (median cumulative dose 12.4 mg) [3].

Previous studies have reported the effects of Teplizumab on beta cell function as measured by stimulated C-peptide [3, 4]. Our goal was to characterize the metabolite profiles of responders versus nonresponders at baseline and to develop a predictive model capable of identifying individuals likely to respond to therapy utilizing baseline metabolite profiles. Using these cohorts, we conducted semi-targeted metabolomics analyses employing liquid chromatography–high-resolution tandem mass spectrometry (LC-HRMS/MS) to identify metabolic signatures associated with treatment response.

## Methods

### Study Design

In the AbATE trial, 52 individuals received Teplizumab and 25 participants received a placebo treatment. Here, we aimed to identify biomarkers for responders vs nonresponders and therefore, our analysis focused on samples available from 41 Teplizumab-treated individuals evaluated at baseline, prior to treatment. Among these participants, 21 individuals did not respond to treatment were classified as “nonresponders”, whereas 20 individuals exhibited a treatment response and were classified as “responders”. Responders were defined as those who experienced less than a 40% decline in baseline C-peptide levels at 2 years from the start of treatment (a threshold exceeded by all placebo participants) [3]. Briefly, this randomized study was conducted at six medical centers and participants were enrolled following informed consent and approval by the institutional review boards at each institution [3].

### Sample Preparation

Serum samples, with minimum freezer thaw, stored at −80°C, were thawed at 4°C. Once thawed, the samples were vortexed, and a volume of 40 μL was pipetted into a microcentrifuge tube. Next, 10 μL internal standard mixture consisting of creatine-D3 H2O (methyl-D3; 4 μg/mL), N-BOC-L-tert-leucine (4 μg/mL), N-BOC-L-aspartic acid (4 μg/mL), succinic acid-2,2,3,3-D4 (4 μg/mL), and salicylic acid (4 μg/mL) was added, along with 250 μL extraction solvent (8/1/1 [v/v/v] acetonitrile/methanol/acetone). Samples were again vortexed then incubated (30 min, 4°C) to further precipitate proteins and centrifuged at 20,000g (10 min, 4°C). The supernatants were transferred and dried (N_2_, 30°C), then stored at −80°C until the day of analysis. On the day of analysis, samples were reconstituted using 40 μL water with 0.1% formic acid, containing BOC-L-tyrosine (2 μg/mL), N-α-BOC-L-tryptophan (2 μg/mL), and BOC-D-phenylalanine (2 μg/mL) as injection standards, vortexed, incubated (15 min, 4°C) and centrifuged at 20,000g (10 min, 4°C). The supernatants were then transferred to liquid chromatography (LC) vials with a fused conical insert [5].

### Liquid Chromatography–High-Resolution Tandem Mass Spectrometry Instrumentation

LC-HRMS/MS analysis was performed using a Vanquish LC system coupled to an Orbitrap Exploris 120 mass spectrometer (Thermo Fisher Scientific). Chromatographic separation was achieved using an ACE Excel 2 C18-PFP (100 × 2.1 mm, 2 μm) column with a mobile phase A consisting of water with 0.1% formic acid and B consisting of acetonitrile. Separation was performed at a flow rate of 0.35 mL/min with gradient elution (0% B to 80% B over 22 minutes). The column temperature was maintained at 25°C. Data were acquired in both positive and negative electrospray ionization modes, with full scan MS performed over an m/z range of 70–1,000 at a resolution of 120,000. Data dependent MS/MS was performed using a normalized collision energy set to 35 and isolation window set to 1.5 m/z, with a mass resolution of 15,000 [5]. Data independent MS/MS was performed using a normalized collision energy set to 30 and isolation m/z range of window 70–700 at a resolution of 60,00.

### Data Processing and Statistical Analysis

Data processing was conducted using MZmine [6] (version 4.5.0), which included mass detection, chromatogram building, smoothing, deconvolution, and library annotation. Full details of the workflow are provided in the electronic supplementary material. Metabolite annotation was conducted using both an in-house library and LC–HRMS/MS spectra from the MassBank of North America to achieve level 2 metabolite identification in both positive and negative ionization modes. All multivariate statistical analyses were performed using MetaboAnalyst [7], where data were sum-normalized, log-transformed, and auto-scaled. Data were not corrected for sex (ESM Fig. 17), age (ESM Fig. 18), or baseline C-peptide (ESM Fig. 19) as the principal component analysis plot showed no distinct clustering. A P-value of <0.05 was selected to identify significant metabolites. All figures were created using the Python programming language.

### Predictive Modeling

To evaluate whether baseline metabolite profiles could predict treatment response to Teplizumab, we developed a supervised Random Forest classifier implemented in Python. This method was selected for its strong performance on categorical outcomes and suitability for high-dimensional, noisy metabolomics data. Additionally, Random Forest is resistant to overfitting in small sample populations [8].

Fifteen metabolites found to be significantly different between responders and nonresponders at baseline (P < 0.05) were used as predictors. To construct the training dataset, 14 responders and 14 nonresponders (representing 70% of the total cohort) were randomly selected using a random fixed seed for reproducibility. The remaining 13 participants were held out as an independent test set. Model development used a pipeline consisting of feature scaling followed by a Random Forest classifier (scikit-learn; n_estimators = 300, random_state = 42). We evaluated generalization using 5-fold stratified cross-validation with area under the curve (AUC) as the primary metric, reporting mean AUC, standard deviation, and 95% confidence intervals, and classifying performance as strong (≥0.80), moderate (≥0.70), or weak (<0.70). Robustness was further evaluated via 1,000-iteration bootstrap resampling for a 95% confidence interval and a 1,000-permutation test for significance to determine whether performance exceeded chance (p < 0.05). Outputs included accuracy, a receiver operating characteristic (ROC) curve with AUC, a confusion matrix, cross-validation metrics, and resampling-based validation statistics.

## Results

Table 1 lists demographic information.

**Table 1.**
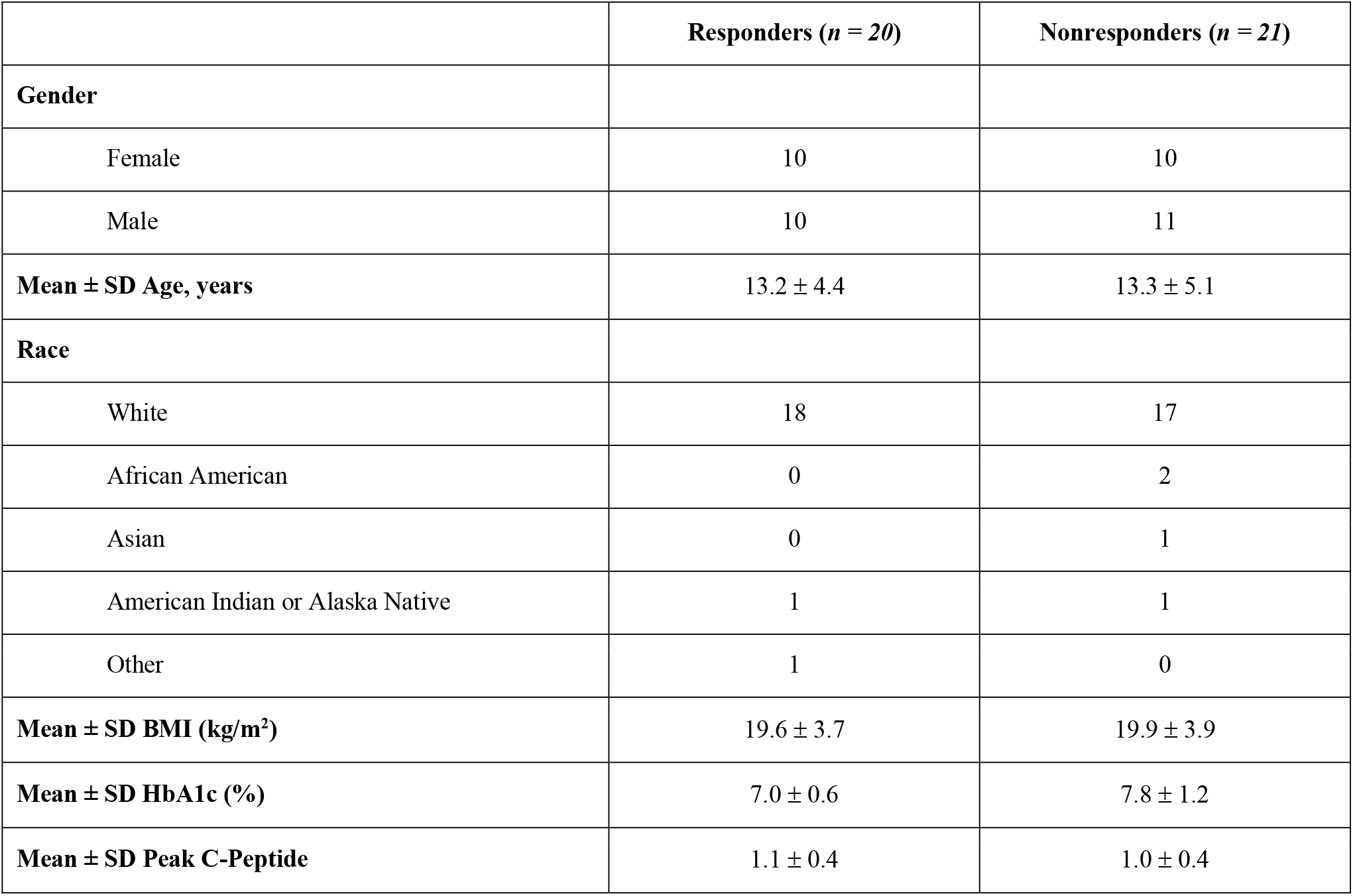
Demographic characteristics of study participants.

### Baseline Metabolite Profiling of Responders versus Nonresponders

Fifteen significant metabolites were identified (Figure 1). Significant metabolites (P < 0.05) included amino acids, their derivatives, and microbial metabolites. At baseline, nonresponders showed higher levels of glutamate and hexanoylcarnitine. In responders, elevated metabolites included TCA cycle intermediates (glutamine, aspartate, malate) and amino acids (lysine). Responders also exhibited higher levels of microbial metabolites (hippurate, indole derivatives, imidazole propionate) and of γ-aminobutyric acid (GABA) and ascorbate.

**Figure 1.**
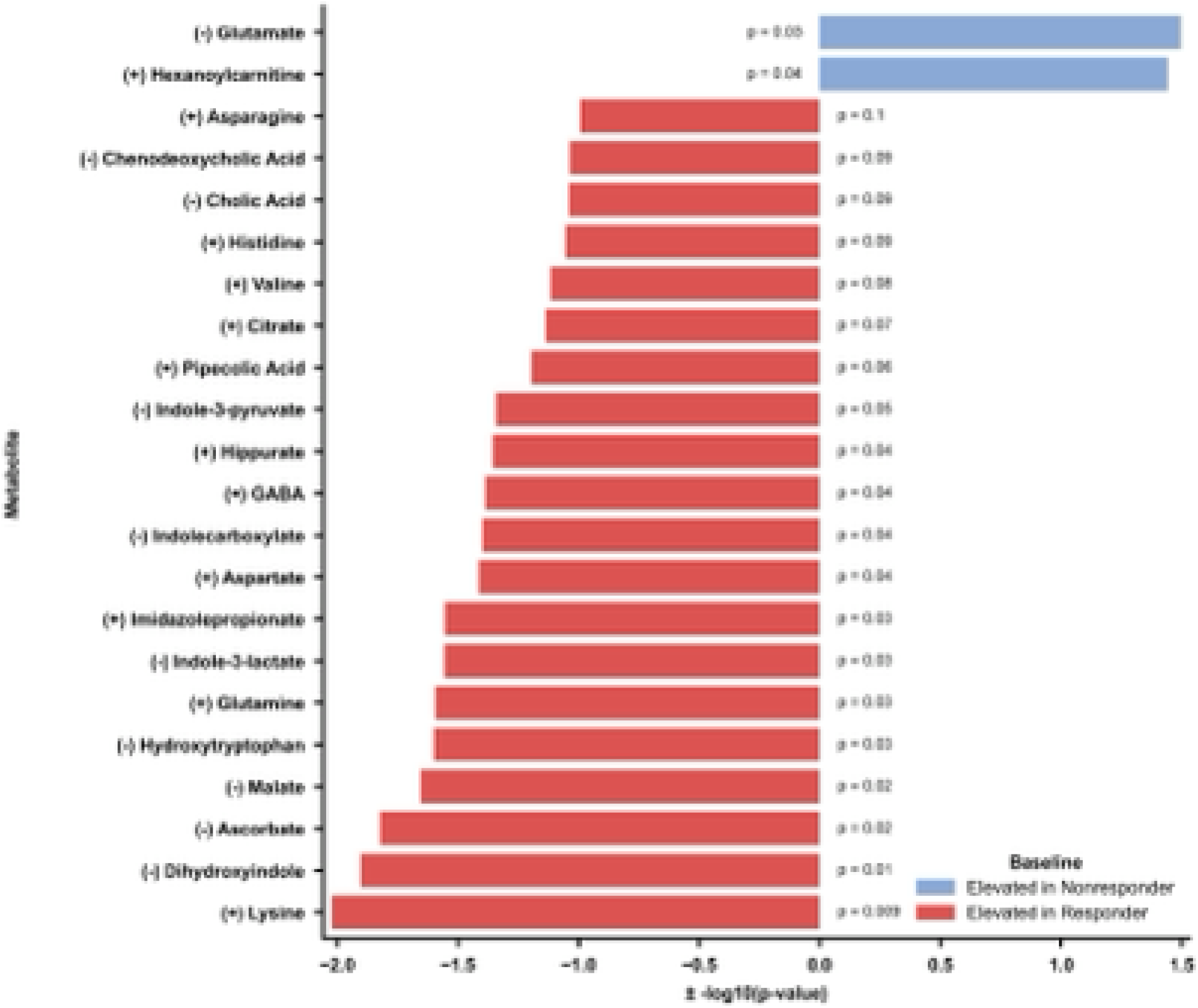
Baseline metabolomic profiling via LC-HRMS/MS. Volcano bar plot illustrating 15 significant metabolites differentiating responders and nonresponders at baseline.

### Predictive Modeling

Using Random Forest, the model was trained to classify samples as either responders or nonresponders. For training, 14 responders and 14 nonresponders were randomly selected from the dataset, while the remaining 13 samples were reserved for testing. The model achieved an accuracy of 0.769. Performance on the test data is summarized in the confusion matrix, where four samples were correctly predicted as responders and six samples as nonresponders (Figure 2A). The ROC curve (Figure 2B) yielded an AUC of 0.881, reflecting high predictive accuracy.

**Figure 2.**
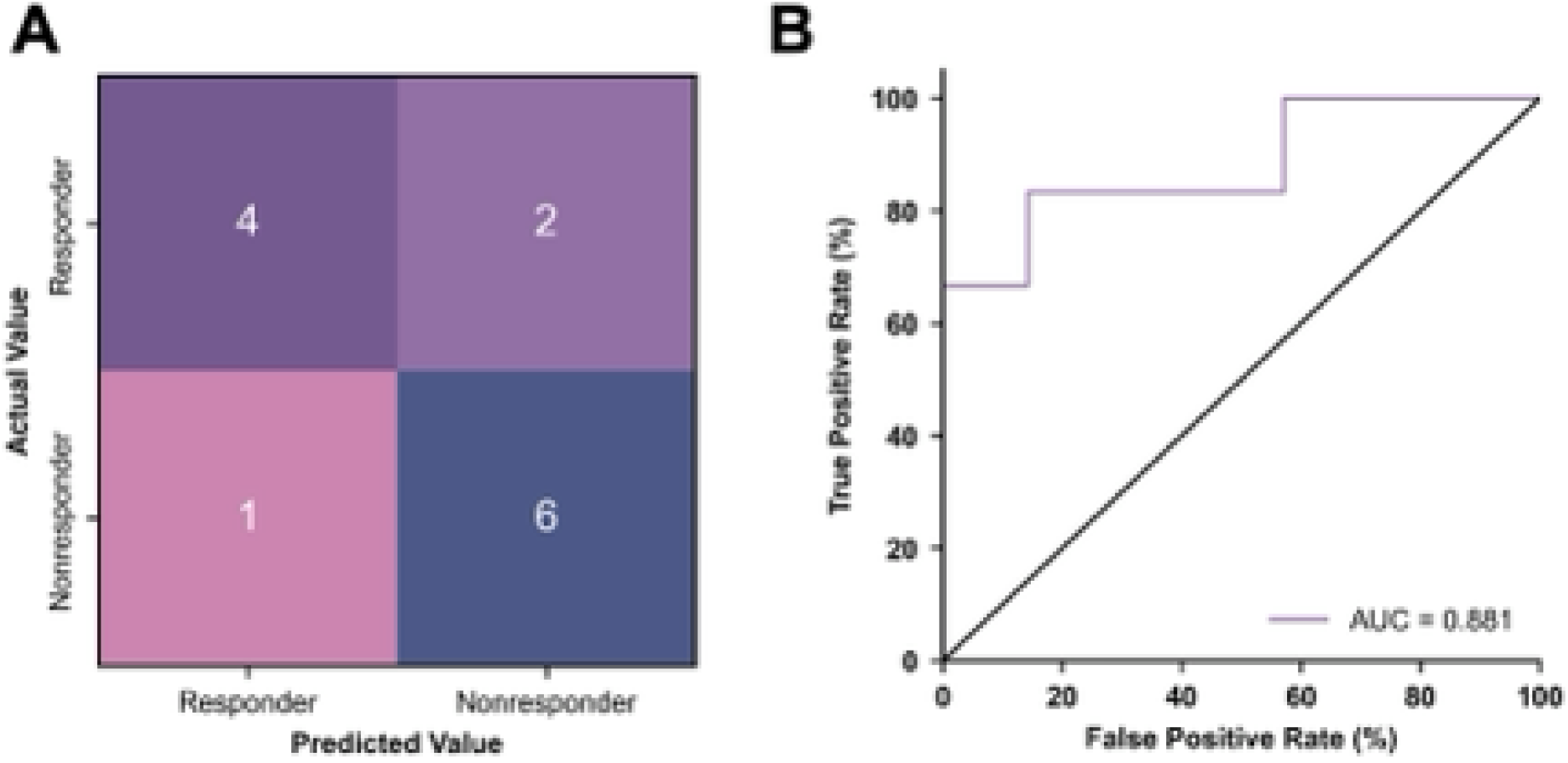
Predictive modeling of treatment response. (A) Confusion matrix summarizing classification performance of the Random Forest model distinguishing responders from nonresponders within the test set (*n* = 13). (B) ROC curve demonstrating high predictive accuracy with an AUC of 0.881.

The Random Forest model demonstrated strong performance during 5-fold cross-validation, yielding fold AUC values ranging from 0.667 to 1.00, with a mean AUC of 0.856 (SD = 0.156) and a 95% confidence interval of 0.719 to 0.992, indicating robust generalization capability. Overfitting assessment revealed a perfect training AUC of 1.000 and a testing AUC of 0.881, producing a modest AUC difference of 0.119, suggesting mild but acceptable overfitting given the small sample size. A bootstrapping analysis (1000 iterations) produced a test AUC confidence interval of 0.619 to 1.000, reflecting moderate stability of the model’s estimates. Finally, the permutation test indicated statistical significance (p = 0.012), confirming that the model performs better than chance. In addition, when using randomized groupings of five of the fifteen significant metabolites, the models demonstrated moderate to weak generalization, and the findings were not statistically significant, indicating that all 15 metabolites are required for prediction.

## Discussion

In this study, we aimed to identify metabolites that differed significantly at baseline between individuals who responded to Teplizumab treatment and those who did not. Fifteen significant metabolites were identified, and a Random Forest accurately classified treatment response based on baseline metabolomic profiles. The model achieved an overall accuracy of 0.769 with an AUC of 0.881, demonstrating that metabolomic profiling at baseline has strong potential to predict therapeutic responsiveness to Teplizumab in individuals with recent-onset T1D.

We observed reduced glutamate levels in individuals who responded to Teplizumab. Glutamate plays a role as a mediator in glutamine-amplified insulin secretion, where glutamate derived from glutamine via glutaminase enhances insulin release through increased Ca^2+^ influx [9]. Elevated glutamate levels, as observed in nonresponders, have been associated with progressive beta cell death [10] and associated with decreased insulin sensitivity and insulin secretion [11]. Therefore, lower glutamate levels at baseline in responders may indicate reduced beta cell stress and preserved insulin secretory function, creating a more favorable metabolic environment for Teplizumab-mediated immune modulation and beta cell preservation. Additionally, we observed elevated levels of glutamine, aspartate, and malate, which are intermediates of the TCA cycle, in individuals who responded to treatment. These elevated TCA cycle metabolites suggest increased mitochondrial activity and enhanced NADPH production, which supports redox balance and beta cell function. NADPH plays a critical role in maintaining redox signaling within beta cells, thereby promoting insulin secretion and cell survival [9]. Consistent with our findings, prior studies have shown that elevated glutamine levels are linked to increased insulin secretion and sensitivity [11]. These findings suggest that responders may begin with stronger beta cell function and metabolic stability, allowing Teplizumab to work more effectively in reducing autoimmune stress on remaining beta cells.

We also observed elevated baseline levels of amino acids in responders, notably lysine. Elevated lysine levels may enhance immune regulation, as lysine has been reported to induce tolerogenic dendritic cells that help maintain immune homeostasis [12]. We also observed differences in gut microbial metabolites, with elevated levels of hippurate, several indole derivatives, and imidazole propionate in responders. Hippurate has been shown in mouse models to improve glucose tolerance and enhance insulin secretion [13]. Elevated hippurate levels at baseline may therefore reflect a healthier gut microbial composition, supporting better beta cell function and glucose regulation prior to treatment. Furthermore, we detected higher levels of several indole derivatives, including indole-3-pyruvate, indole carboxylate, indole-3-lactate, hydroxytryptophan, and dihydroxyindole. These microbial metabolites, derived from tryptophan catabolism activate the aryl hydrocarbon receptor, which promotes regulatory T cell differentiation and supports immune tolerance [14]. Elevated imidazole propionate, a histidine-derived metabolite, may reflect an activated immune state and systemic inflammation [15], potentially enhancing responsiveness to Teplizumab’s immune-modulating effects. Although not statistically significant (P = 0.09), histidine levels were higher in responders, suggesting that greater baseline histidine availability could contribute to elevated imidazole propionate. Consequently, elevated levels of these metabolites at baseline may reflect a gut microbiome composition that favors enhanced insulin secretion and regulatory immune pathways, potentially facilitating a more effective immunomodulatory response to Teplizumab.

Finally, we observed elevated baseline levels of GABA and ascorbate in individuals who responded to Teplizumab. GABA is synthesized and secreted by pancreatic beta cells, where it plays a critical role in modulating insulin release and immune signaling within the islet microenvironment. In T1D, progressive beta cell loss leads to reduced GABA production [16]. Thus, elevated baseline GABA may mark preserved beta cell mass and function, creating favorable conditions for Teplizumab. Ascorbate, or vitamin C, functions as an antioxidant that protects immune cells from oxidative stress and supports proper T-cell differentiation and activation [17]. Elevated ascorbate levels in responders may reflect a more balanced redox state and a healthier immune profile, promoting immune tolerance. In contrast, we observed elevated levels of hexanoylcarnitine, a medium-chain acylcarnitine, in nonresponders. Elevated medium-chain acylcarnitines have been associated with mitochondrial stress and impaired beta cell function [18]. These findings suggest that nonresponders may have greater underlying beta cell dysfunction at baseline, reducing the potential benefit of Teplizumab’s immune-modulating and beta cell preserving effects.

A limitation of our study is the relatively small sample size, which may affect generalizability. However, the Random Forest model demonstrated strong predictive performance despite this limitation, achieving an accuracy of 0.769 and an AUC of 0.881. While validation in larger cohorts is needed, the Random Forest model offers advantages in reducing overfitting and enhancing feature selection, supporting its potential for identifying predictive metabolic signatures. Additionally, our analysis focused exclusively on Teplizumab, a single immune therapy.

Overall, the baseline metabolomic profile of Teplizumab responders indicates preserved beta cell function, enhanced mitochondrial activity, and immune tolerance. Elevated amino acid, TCA cycle, and microbial metabolites suggest active mitochondrial metabolism and regulatory immune signaling, while lower glutamate and acylcarnitine levels indicate reduced metabolic stress. Together, these findings may provide insight into why certain individuals respond more favorably to Teplizumab and suggest that baseline metabolite profiles could be used to predict treatment response. Future studies should expand this approach to include other immunotherapies and examine both baseline and post-treatment metabolomic profiles.

## Supporting information

ESM

## Data Availability

The data sets generated and analyzed are available upon reasonable request.

## Abbreviations

T1D: Type 1 diabetes
AbATE: Autoimmunity-Blocking Antibody for Tolerance in Recently Diagnosed Type 1 Diabetes
TCA: Tricarboxylic acid
LC-HRMS/MS: liquid chromatography–high-resolution tandem mass spectrometry
LC: liquid chromatography
ROC: receiver operating characteristic
AUC: area under the curve
GABA: γ-aminobutyric acid

## Data and Resource Availability

The data sets generated and analyzed are available upon reasonable request.

## Authors’ Relationships and Activities

L.E.H. consulted for GenEdit, Inc. S.S. served on a scientific advisory board for Sanofi.

## Contribution Statement

E.R.F. performed sample analysis, data analysis and interpretation, and wrote the manuscript. L.E.H., S.S., H.M.I., and T.J.G. provided oversight of the research, reviewed all data and findings, and edited the manuscript. T.J.G. is the guarantor of this work and, as such, had full access to all the data in the study and takes responsibility for the integrity of the data and the accuracy of the data analysis.

## Graphical Abstract

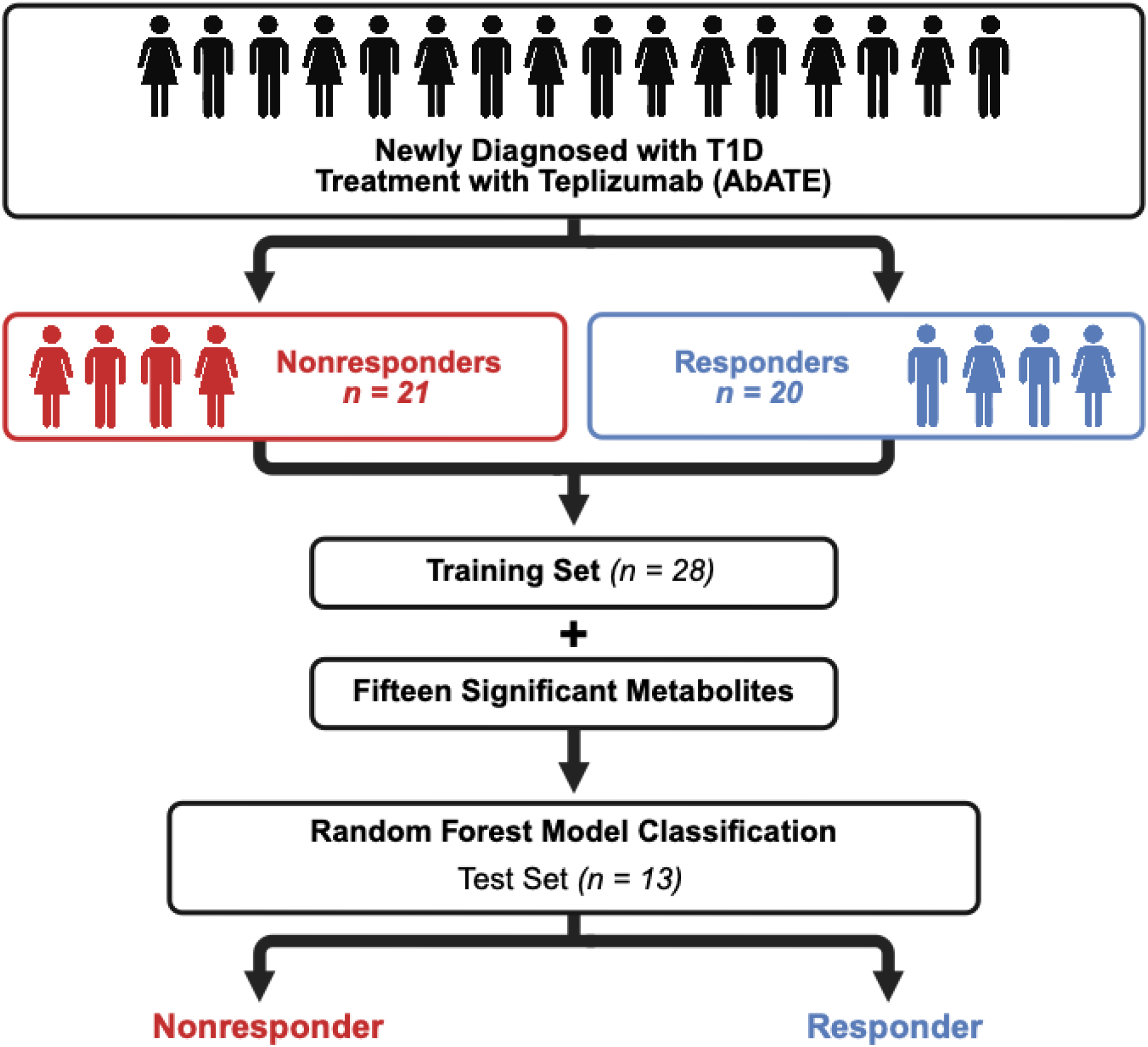

