## Supplementary material for "Baseline Serum Metabolites as Predictors of Teplizumab Response in Individuals with Type 1 Diabetes": ESM

**<u>Electronic supplementary material</u>**

### Electronic supplementary figures


**ESM Fig 1.** MZmine 4.5.0 batch queue used. Page 2

**ESM Fig 2.** MZmine 4.5.0 mass detection parameters used (MS1). Page 3

**ESM Fig 3.** MZmine 4.5.0 mass detection parameters used (MS2). Page 4

**ESM Fig 4.** MZmine 4.5.0 chromatogram builder parameters used. Page 5

**ESM Fig 5.** MZmine 4.5.0 smoothing parameters used. Page 6

**ESM Fig 6.** MZmine 4.5.0 local minimum feature resolver parameters used. Page 7

**ESM Fig 7.** MZmine 4.5.0 13C isotope filter parameters used. Page 8

**ESM Fig 8.** MZmine 4.5.0 isotopic peaks finder parameters used. Page 9

**ESM Fig 9.** MZmine 4.5.0 join aligner parameters used. Page 10

**ESM Fig 10.** MZmine 4.5.0 feature list rows filter parameters used. Page 11

**ESM Fig 11.** MZmine 4.5.0 feature finder parameters used. Page 12

**ESM Fig 12.** MZmine 4.5.0 duplicate peak filter parameters used. Page 13

**ESM Fig 13.** MZmine 4.5.0 correlation grouping parameters used. Page 14

**ESM Fig 14.** MZmine 4.5.0 spectral library search parameters used. Page 15

**ESM Fig 15.** MZmine 4.5.0 lipid annotation parameters used. Page 16

**ESM Fig 16.** MZmine 4.5.0 spectral / molecular networking parameters used. Page 17

**ESM Fig. 17** Principal Component Analysis by Gender. Page 18

**ESM Fig. 18** Principal Component Analysis by Age. Page 19

**ESM Fig. 19** Principal Component Analysis by Baseline C-peptide. Page 20

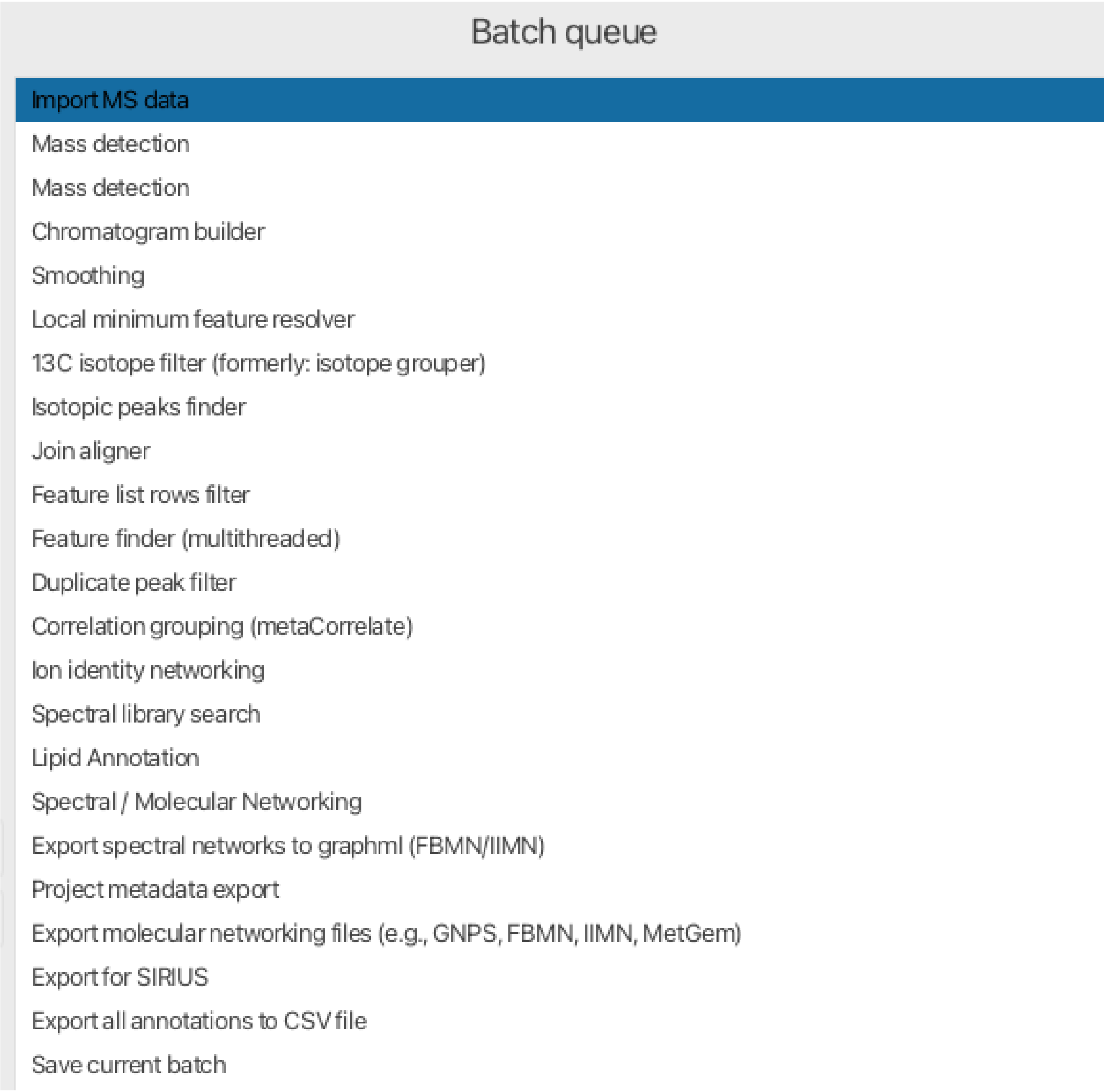


**ESM Fig 1.** MZmine 4.5.0 batch queue used.

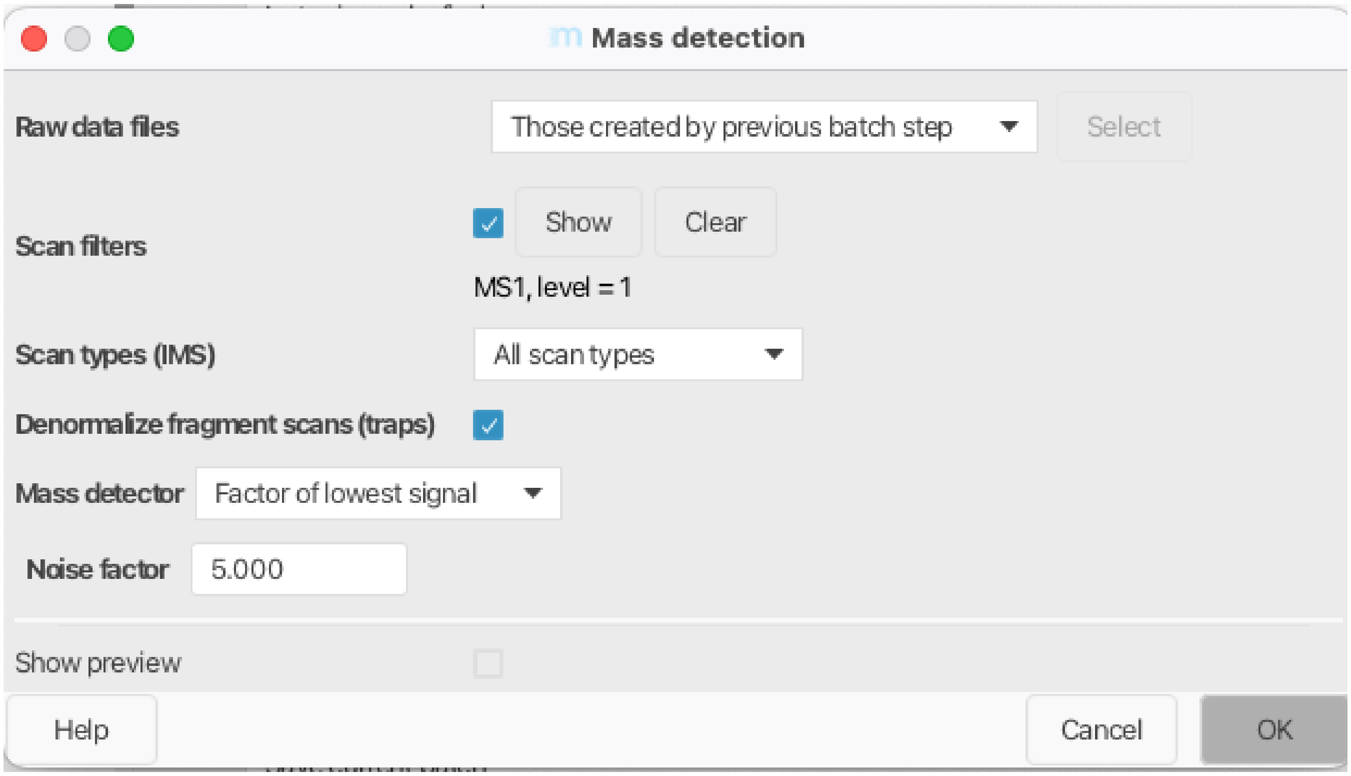


**ESM Fig 2.** MZmine 4.5.0 mass detection parameters used (MS1).

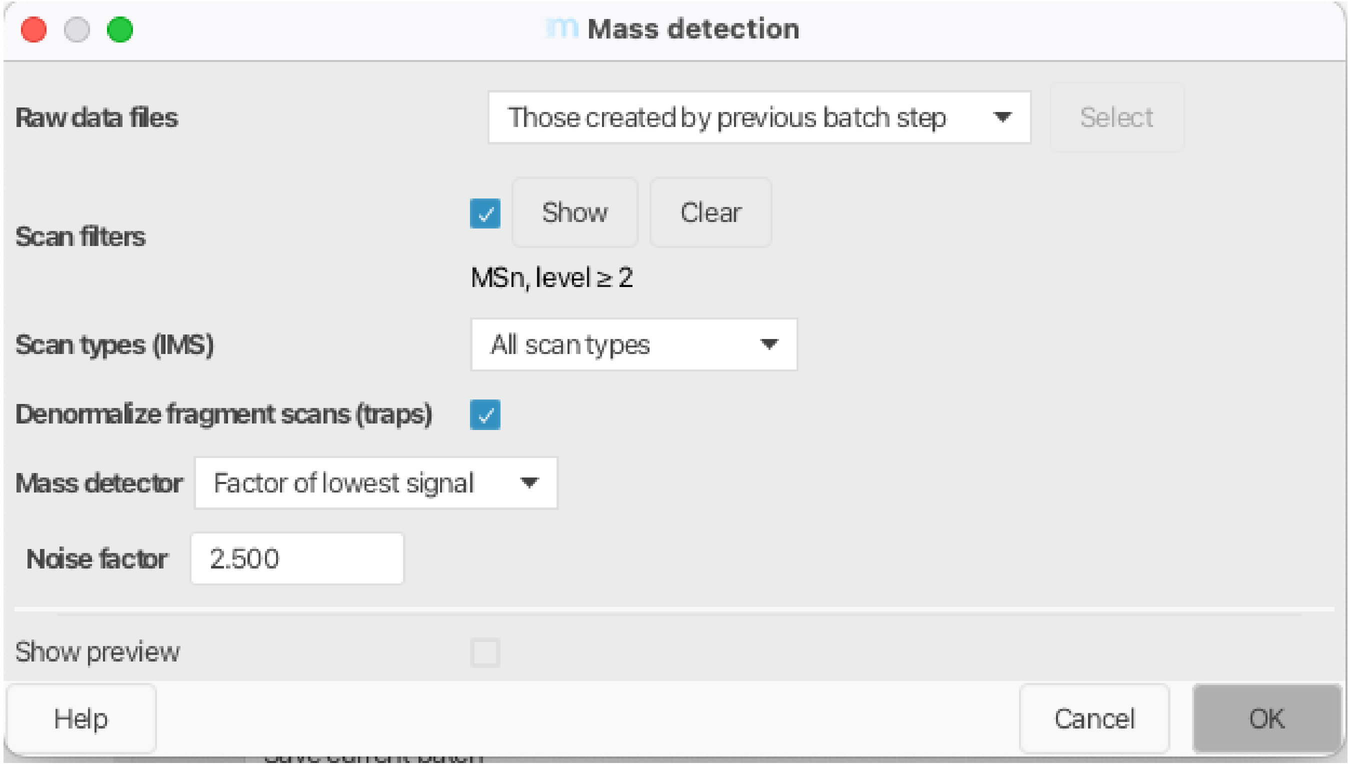


**ESM Fig 3.** MZmine 4.5.0 mass detection parameters used (MS2).

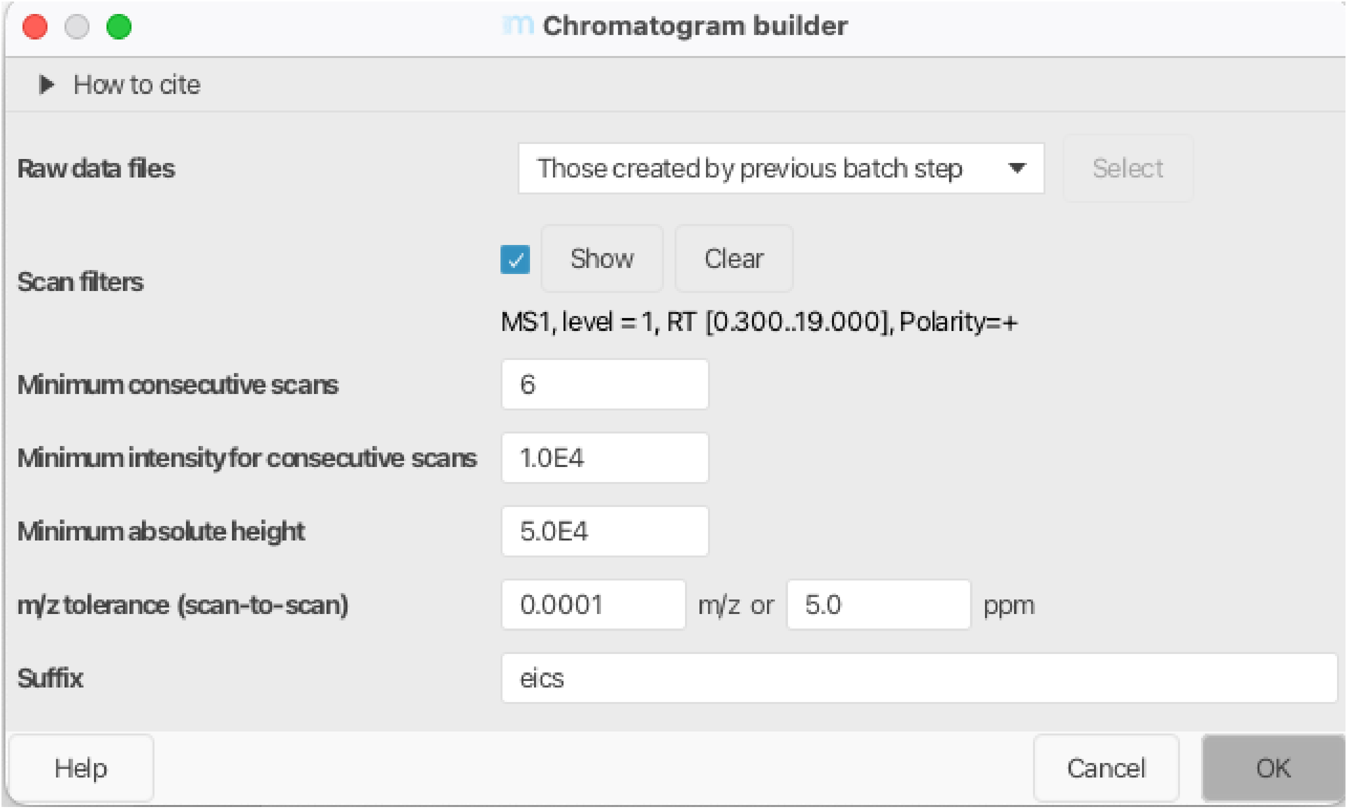


**ESM Fig 4.** MZmine 4.5.0 chromatogram builder parameters used.

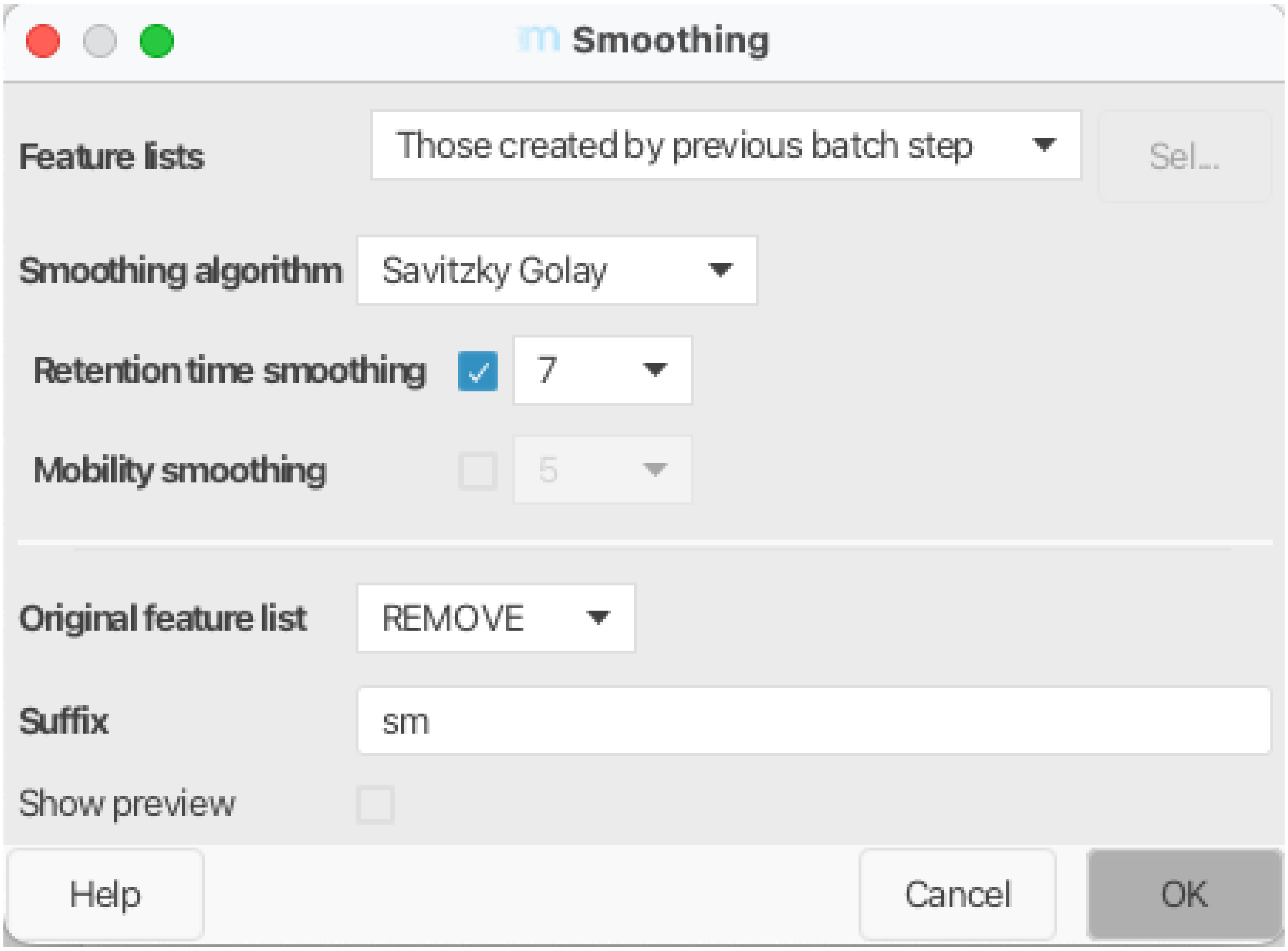


**ESM Fig 5.** MZmine 4.5.0 smoothing parameters used.

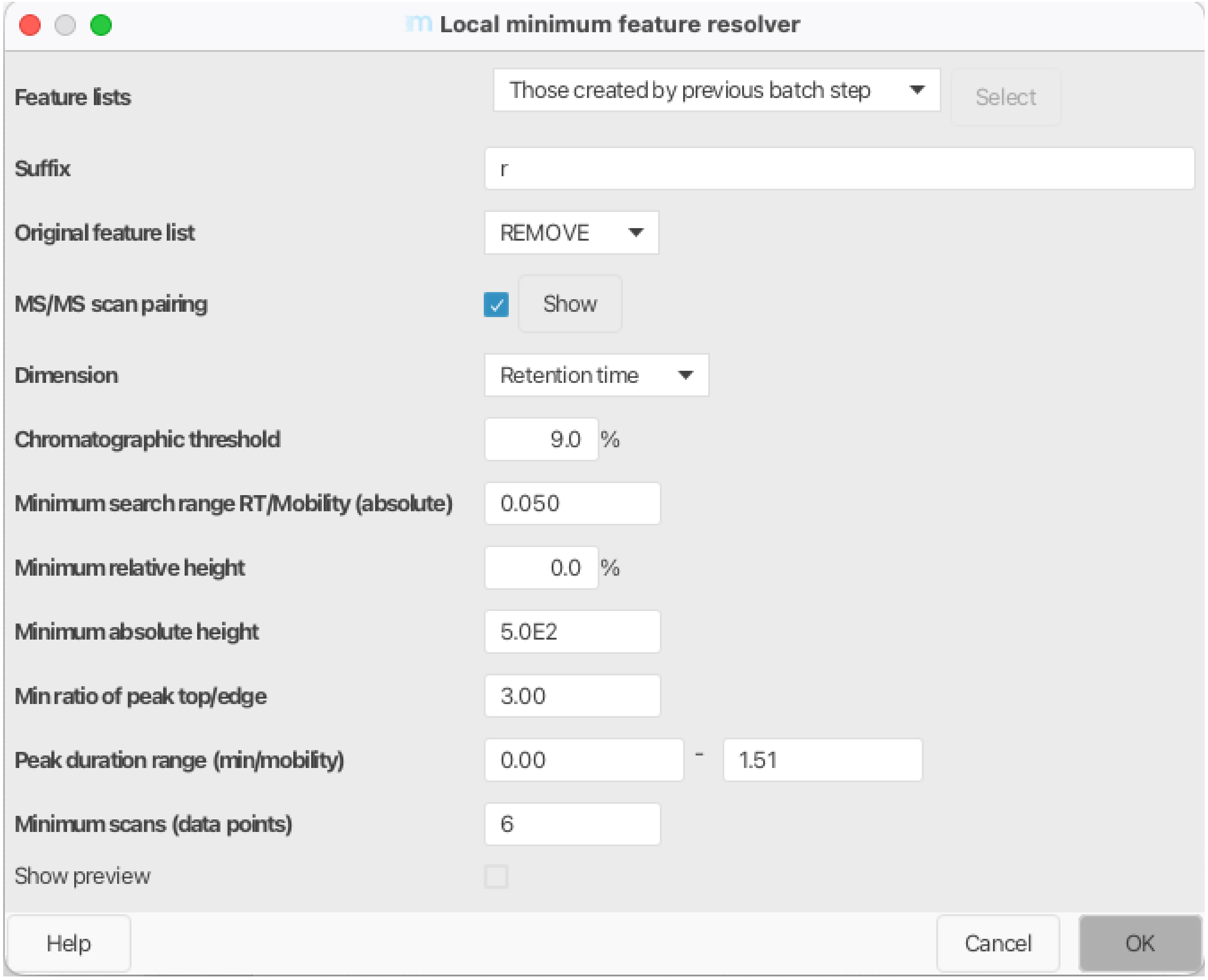


**ESM Fig 6.** MZmine 4.5.0 local minimum feature resolver parameters used.

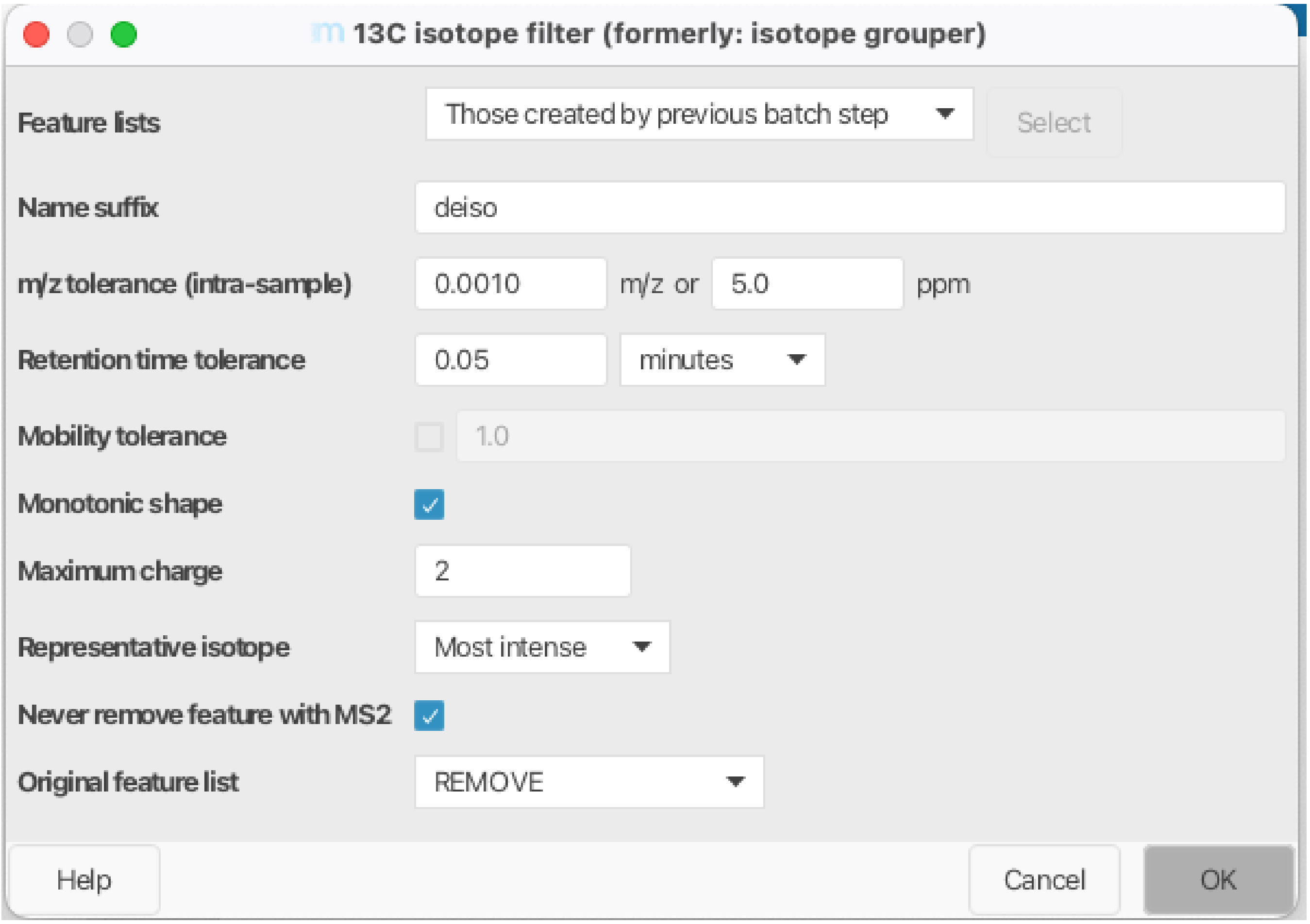


**ESM Fig 7.** MZmine 4.5.0 13C isotope filter parameters used.

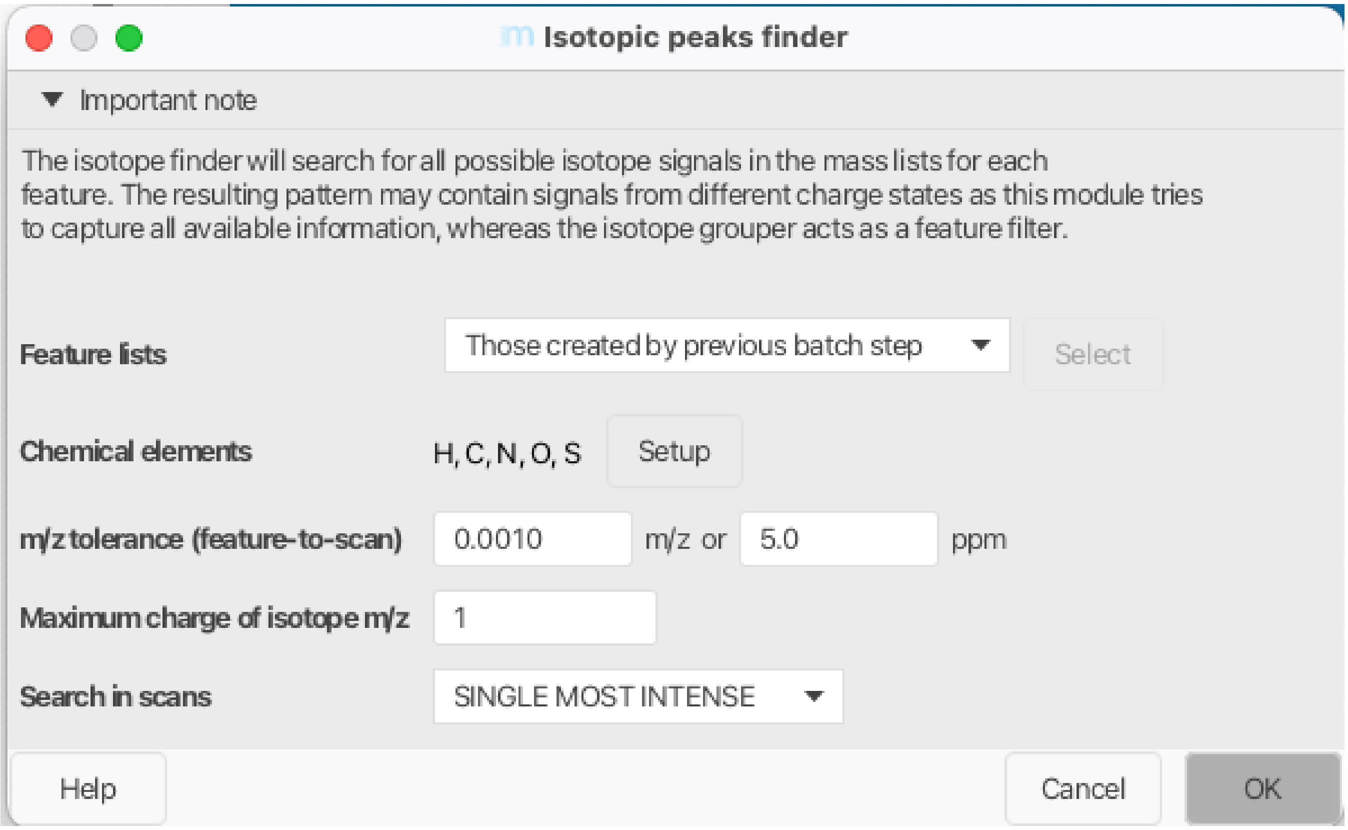


**ESM Fig 8.** MZmine 4.5.0 isotopic peaks finder parameters used.

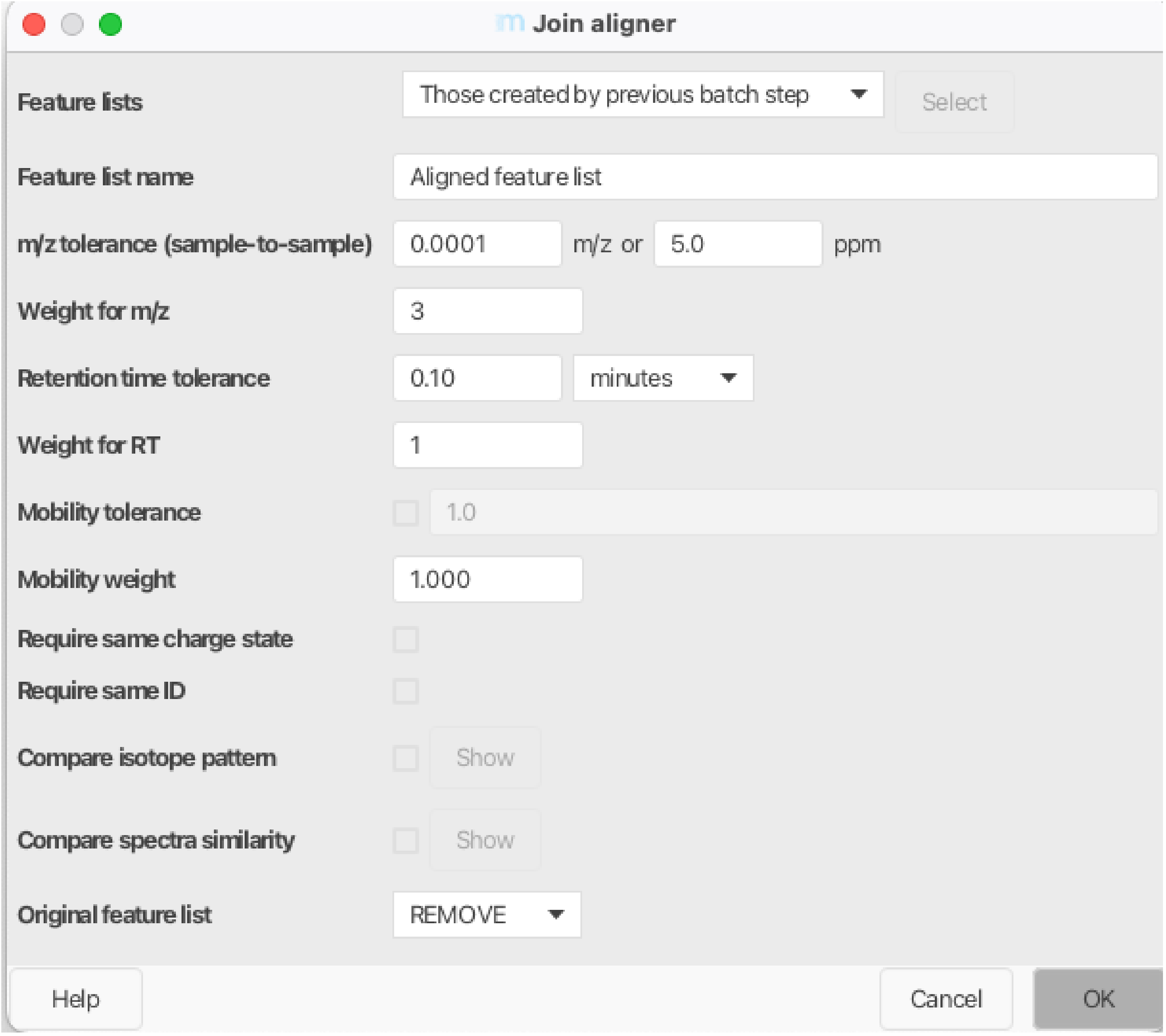


**ESM Fig 9.** MZmine 4.5.0 join aligner parameters used.

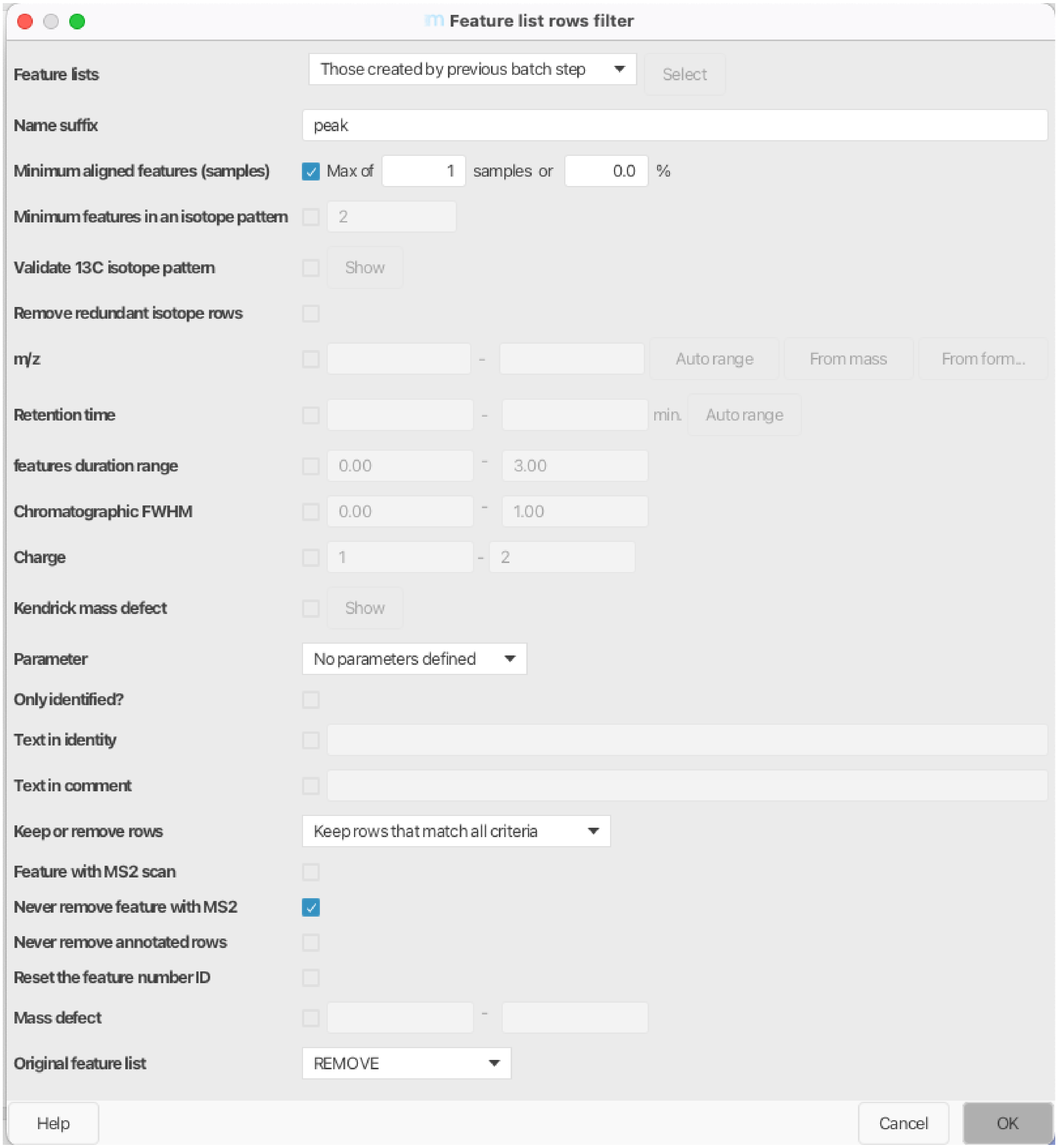


**ESM Fig 10.** MZmine 4.5.0 feature list rows filter parameters used.

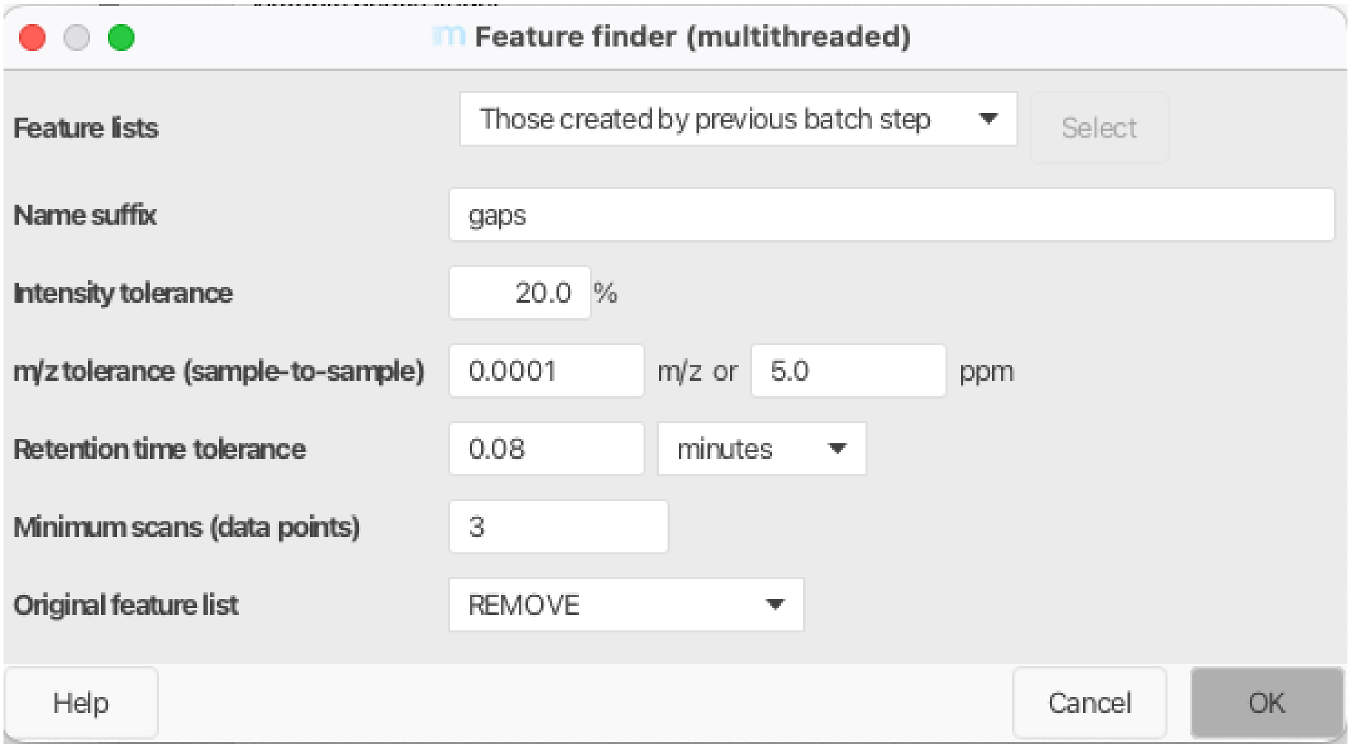


**ESM Fig 11.** MZmine 4.5.0 feature finder parameters used.

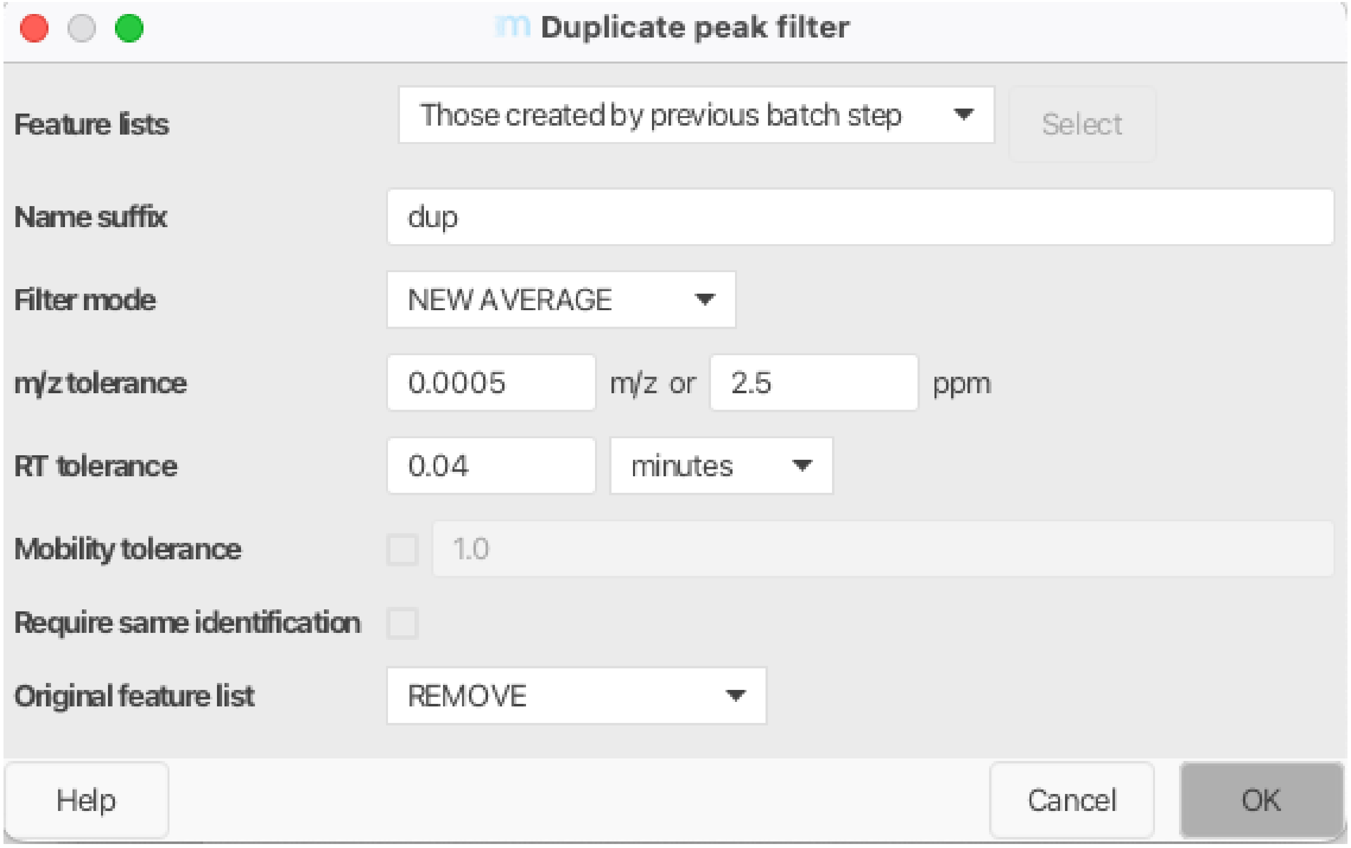


**ESM Fig 12.** MZmine 4.5.0 duplicate peak filter parameters used.

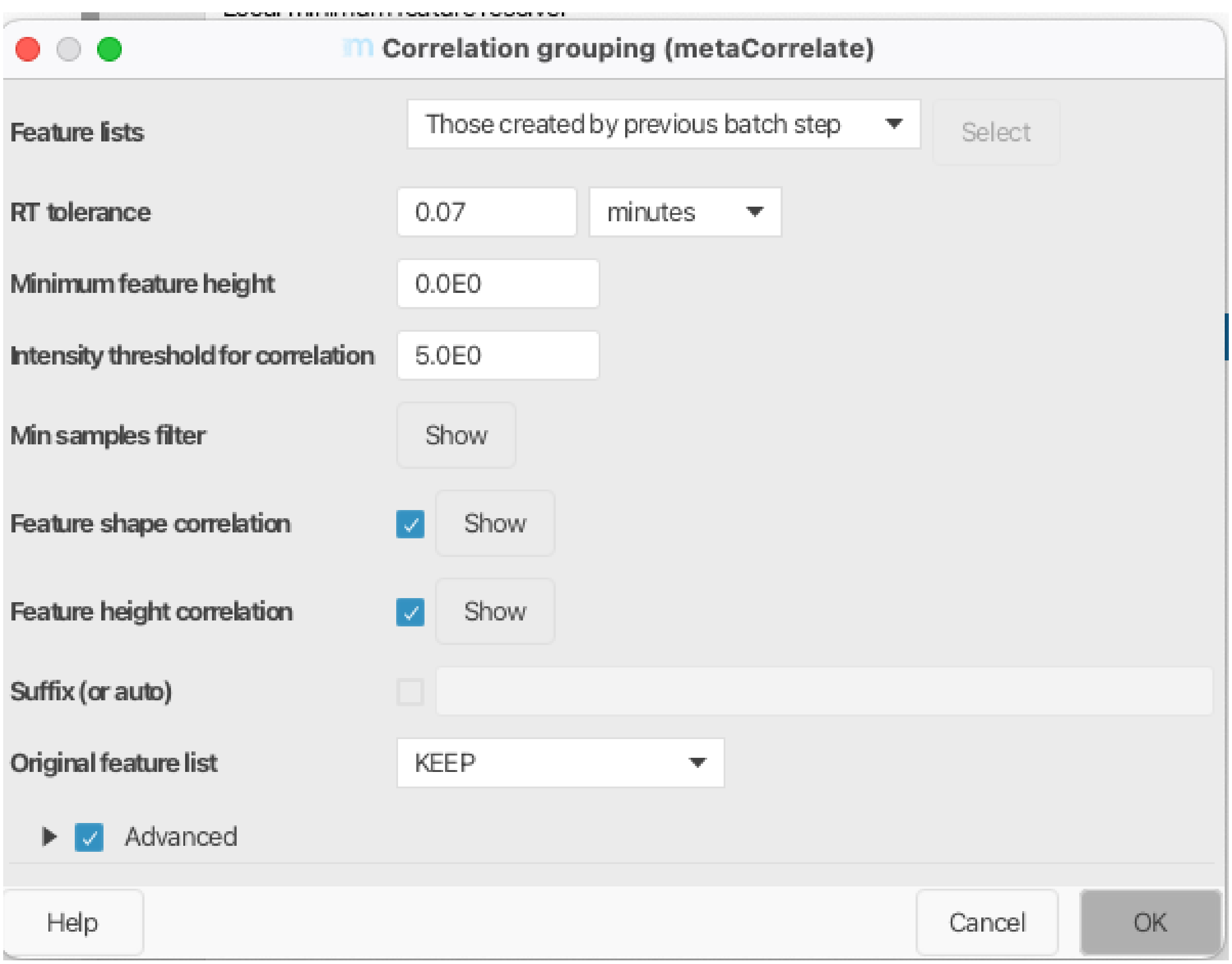


**ESM Fig 13.** MZmine 4.5.0 correlation grouping parameters used.

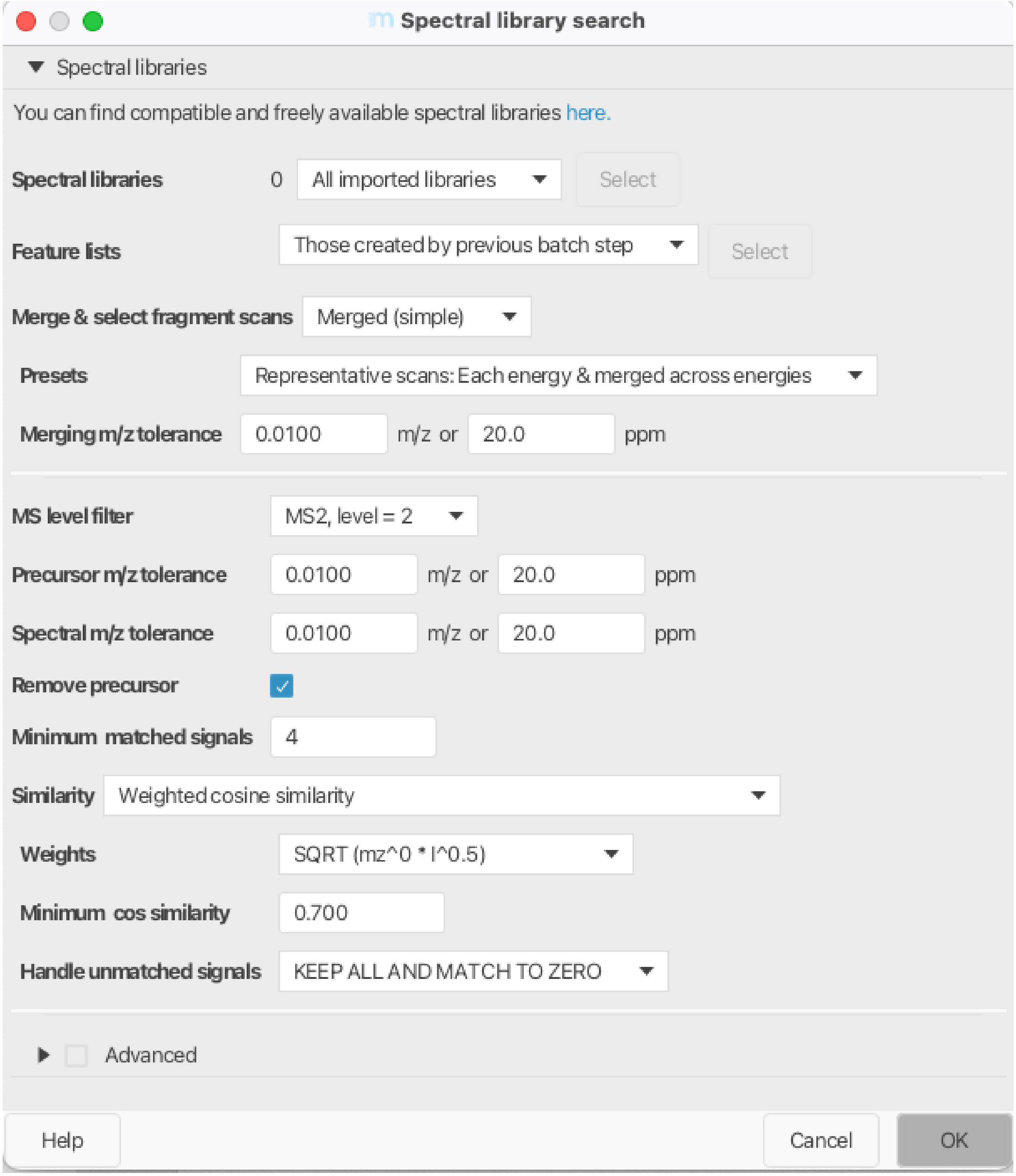


**ESM Fig 14.** MZmine 4.5.0 spectral library search parameters used.

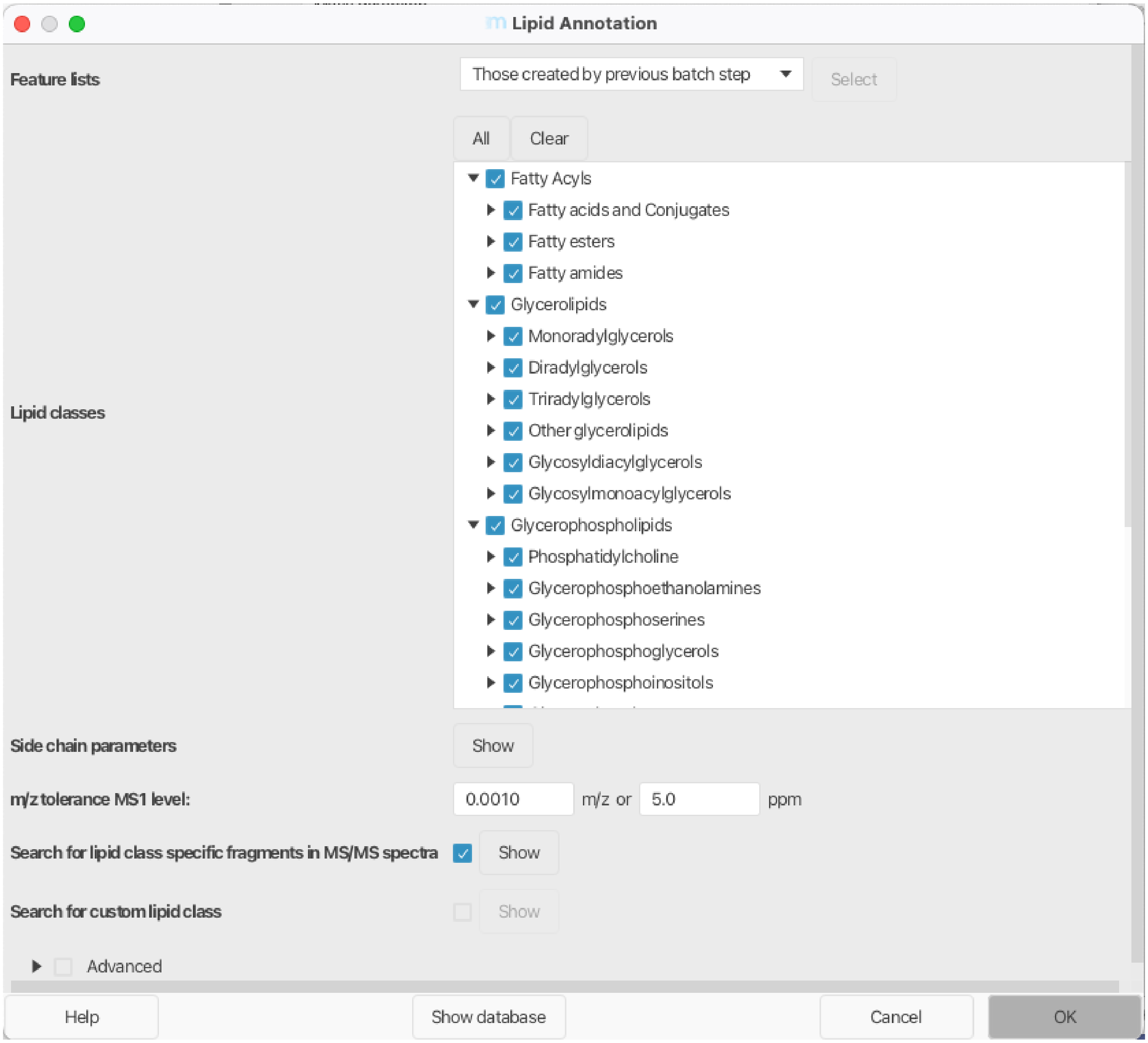


**ESM Fig 15.** MZmine 4.5.0 lipid annotation parameters used.

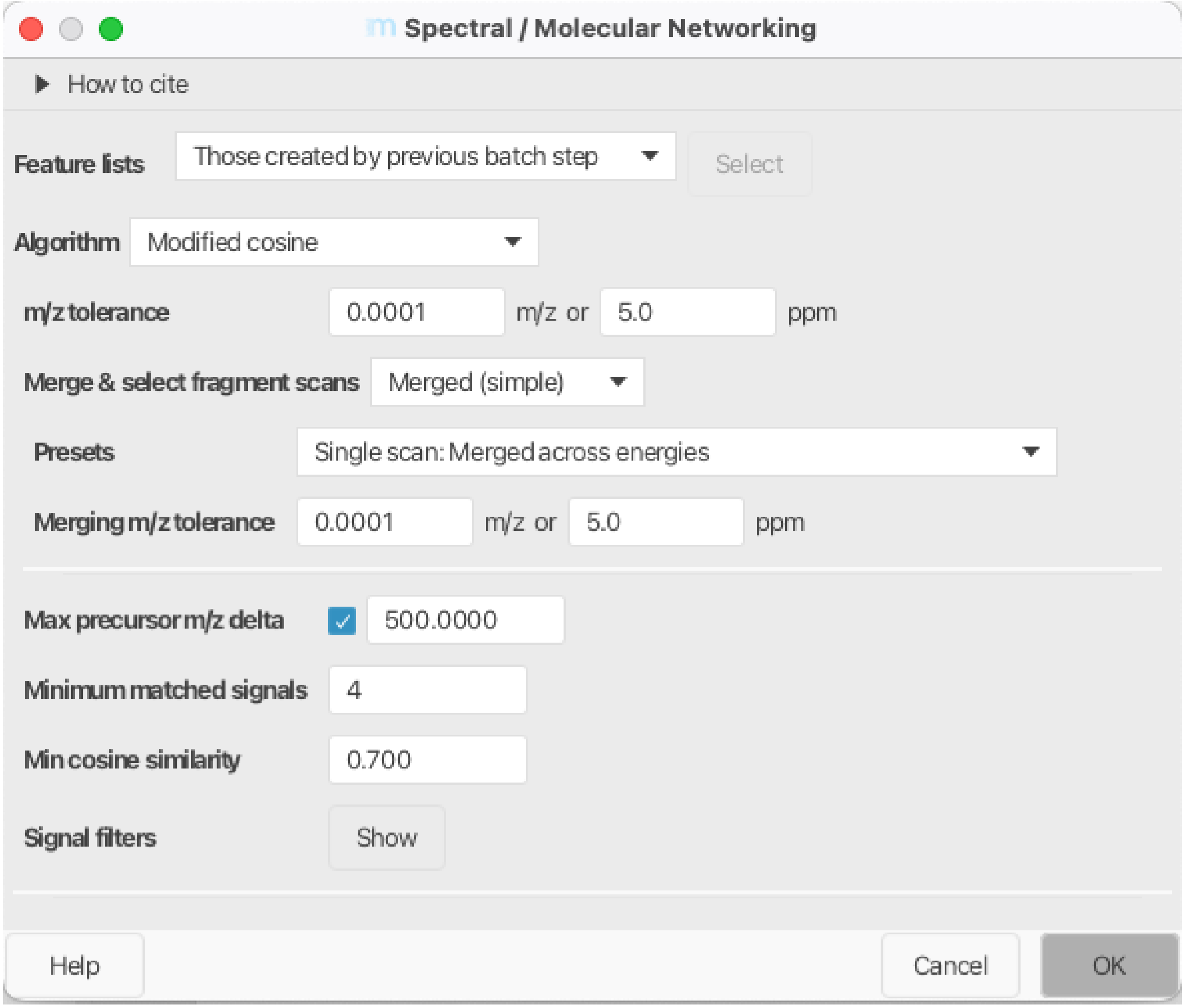


**ESM Fig 16.** MZmine 4.5.0 spectral / molecular networking parameters used.

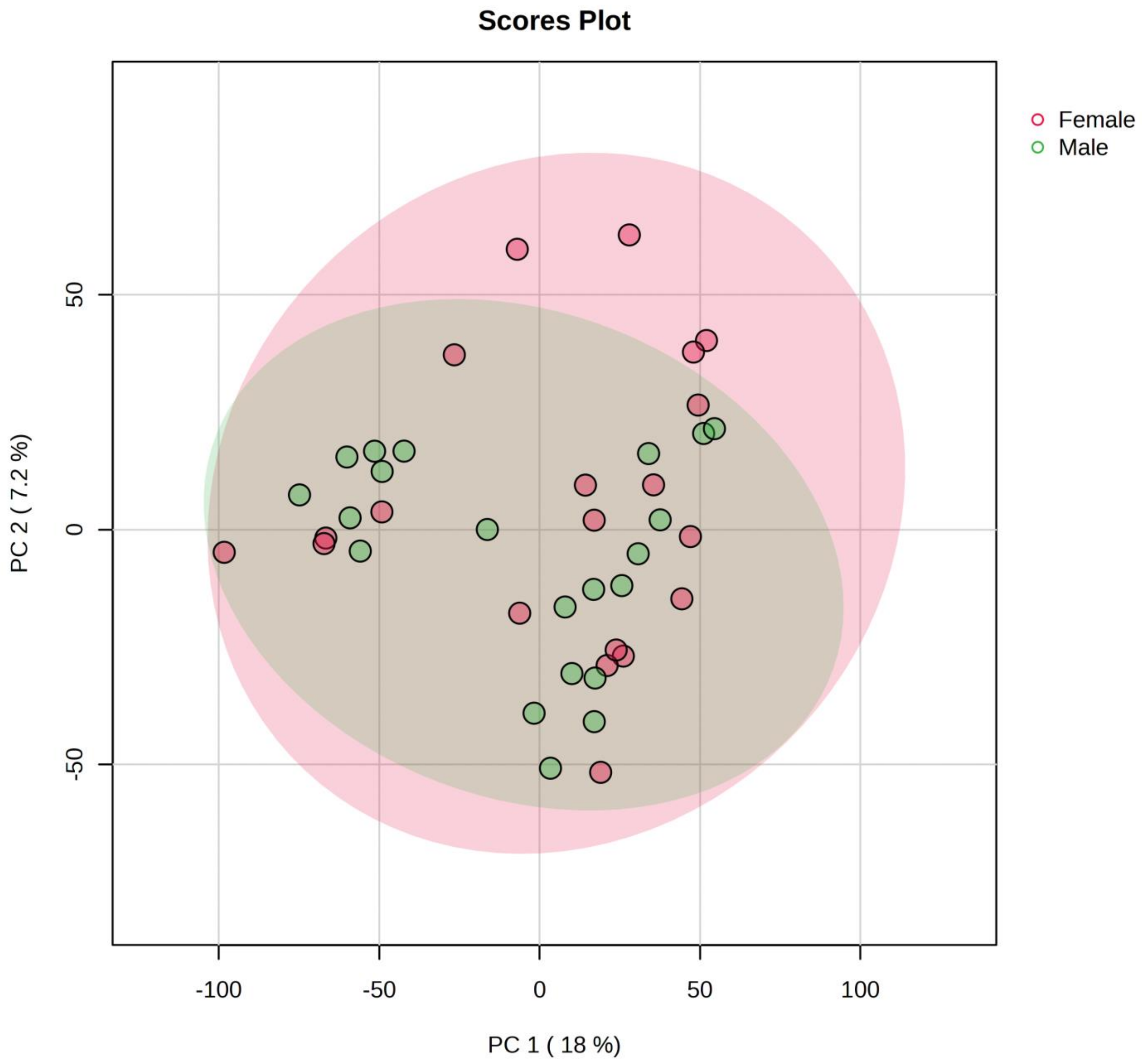


**ESM Fig 17.** Principal Component Analysis by Gender. No distinct clustering by gender.

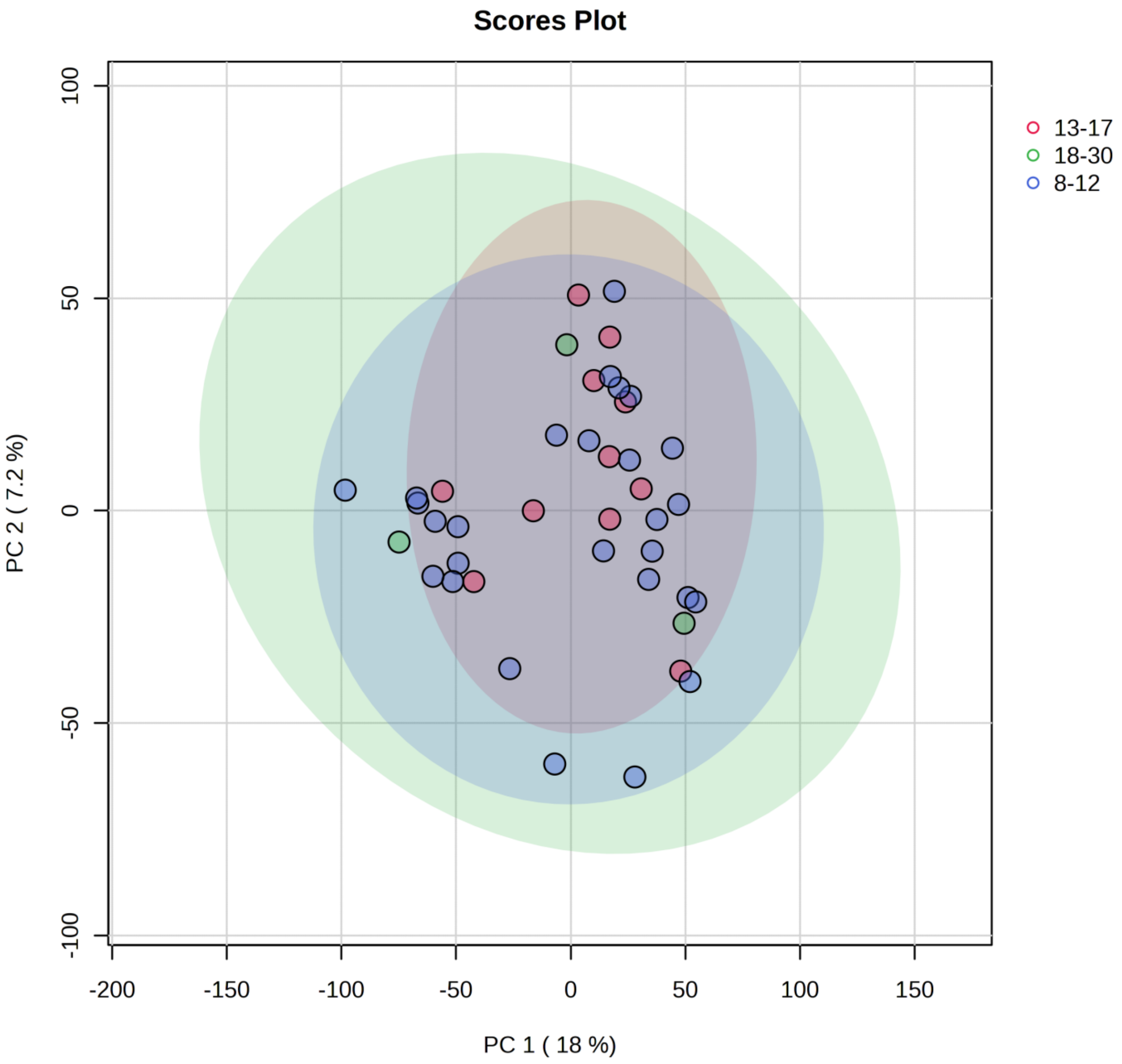


**ESM Fig 18.** Principal Component Analysis by Age. No distinct clustering by age grouping.

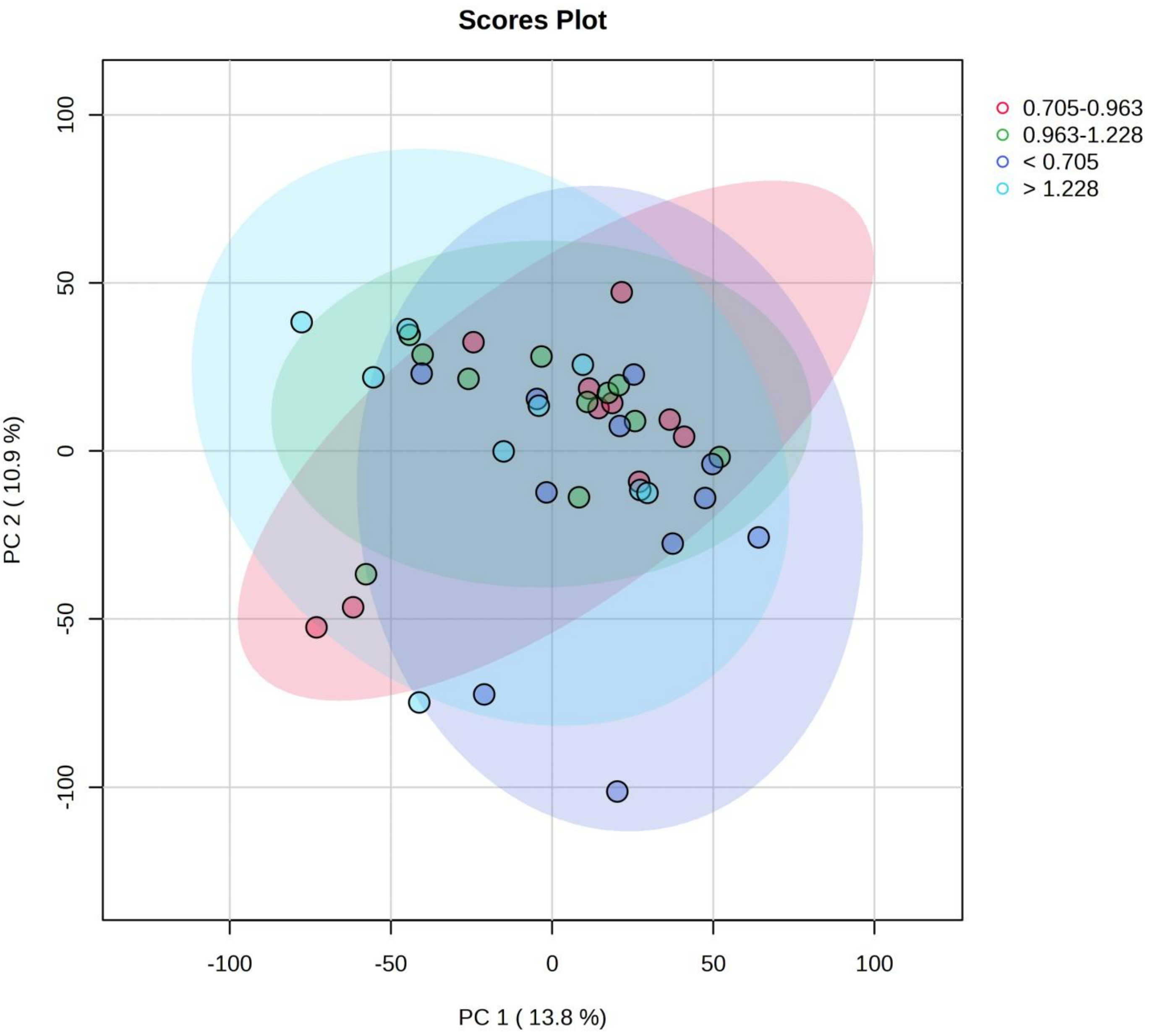


**ESM Fig. 19** Principal Component Analysis by Baseline C-peptide. No distinct clustering by baseline C-peptide grouping.
